# The missing link: Electronic health record linkage across species offers opportunities for improving One Health

**DOI:** 10.1101/2025.03.25.25324490

**Authors:** Kathleen R. Mullen, Nadia Saklou, Adam Kiehl, Toan C. Ong, G. Joseph Strecker, Sabrina Toro, Sue VandeWoude, Ian M. Brooks, Tracy Webb, Melissa A. Haendel

## Abstract

**Objective:** Significant opportunities for understanding the co-occurrence of conditions across species in coincident households remain untapped. We determined the feasibility of creating a Companion Care Registry (CCR) for analysis of health data from the University of Colorado Health (UCHealth) patients and their companion animals who received veterinary care at the geographically-adjacent Colorado State University Veterinary Teaching Hospital (CSU-VTH).

**Materials and Methods:** Using a hybrid deterministic and probabilistic record linkage method, non-medical Personally Identifiable Information was securely matched to determine the total number of UCHealth patients within the HIPAA-compliant Health Data Compass Research Data Warehouse (2015-2024) who took a companion animal to the CSU-VTH (2019-2024).

**Results:** 12,115 matches were identified, indicating 29% of CSU-VTH clients were UCHealth patients.

**Discussion:** The overlap between CSU-VTH clients and UCHealth patients underscores the potential feasibility and utility of a CCR.

**Conclusion:** This work provides a mechanism to evaluate environmental and inter-species influences on One Health.

## BACKGROUND AND SIGNIFICANCE

Accelerating precision medicine within the context of the learning healthcare system requires shifting how we structure, share, and collaboratively analyze data in clinical and translational science. Advances in connecting and integrating information across human health contexts have historically moved relatively slowly. Data from veterinary patients (e.g., veterinary electronic health records (vEHR)) offer an underutilized resource for accelerating translational science and precision medicine. For context, 66% percent or 86.9 million U.S. families own a companion animal, and over 85% of owners consider their pet(s) part of the family.[1,2] There is growing awareness of the relevancy of cross-species data, the importance of the human-animal bond, and the need for One Health approaches to health problems. One Health is an international effort that recognizes the interdependence of human, animal, and environmental health. Accordingly, exploring shared veterinary and human health data has the potential to positively impact patient outcomes.[3]

Many disease definitions and proposed pathophysiologies are similar across species, highlighting their potential translational relevance. Because people and their companion animals coexist in the same household (dogs and cats sharing the indoor and/or outdoor environment and horses sharing the outdoor environment with humans), animals may help uncover risk factors, disease pathways, and novel interventions for shared diseases. For example, the body mass index of adult dog owners and dog body condition score were positively correlated in one study, where nearly 58% of dog owners and 63% of dogs were overweight or obese[4], highlighting the cross-species prevalence and need for successful intervention strategies. Shared behaviors among animal owners and their companion animals suggest that improving the health of one species may have indirect health benefits to the other. Dog ownership was reviewed as a solution to promote exercise among adults with prediabetes and Type 2 diabetes (T2D), but more research is needed to fully evaluate the impact of dog ownership on cardiovascular disease risk in patients with T2D.[5] In addition, household environment potentially has significant health impacts, such as through exposure to air and water pollution and endocrine disrupting compounds.[6–9] In one study, dog owners in more disadvantaged neighborhoods reported less on-leash walking activity compared to owners in advantaged neighborhoods[10], suggesting that companion animal data can potentially fill significant gaps in understanding health disparities.

## OBJECTIVE

We hypothesize that an Observational Medical Outcomes Partnership (OMOP) Common Data Model (CDM)-based[11] Companion Care Registry (CCR) for geographically overlapping human and veterinary medical institutions would enable investigation of the co-occurrence of conditions and environmental influences in humans and companion animals in coincident households. This project sought to assess the feasibility, in terms of available data, technology, and governance of creating a CCR to assess human and veterinary medical and environmental data at a household level to ultimately improve human and animal patient outcomes.

## MATERIALS AND METHODS

In this feasibility study, de-identified aggregate clinical data from the CSU-VTH vEHR and from the UCHealth Health Data Compass (HDC) research data warehouse (RDW)[12] at the University of Colorado Anschutz (CUA) were compared to determine similarities and differences between the veterinary and human medical datasets. Cell counts <10 were suppressed.

Additionally, the number of clients of the CSU-VTH who were also patients of UCHealth was determined. Non-medical Personally Identifying Information (PII) from the CSU-VTH client records from years 2019-2024 was linked with patient records in the HDC RDW from years 2015-2024. Exemption status was obtained from the Colorado Multiple Institutional Review Board for the study (“Pilot Project to Assess Overlap between People Receiving Healthcare Locally (UCHealth) and Companion Animal Care at the CSU VTH” (COMIRB # 23-0940)) and deemed not human subjects research by the CSU Institutional Review Board (CSU IRB #1920). A data use agreement was developed and signed by the CSU Board of Governors and the Regents of the University of Colorado for and on behalf of CUA agreeing to the scope and purpose of the project and the obligations and activities of the data recipient, CUA.

PII (first name, last name, street address, city, 5-digit zip code, phone number and email address) from the CSU-VTH client records was delivered to the Google Cloud-based HDC RDW at CUA via encrypted file transfer to a HIPAA-compliant virtual machine (VM). CSU PII was matched to patient records in the RDW using a hybrid deterministic and probabilistic record linkage method contained in the CU Record Linkage (CURL) Python package (**Table 1**).[13]

**Table 1:** Deterministic and probabilistic record linkage method.

| Method | Linkage variable |
| --- | --- |
| Deterministic 1 | First6ofFN + First6ofLN + phone |
| Deterministic 2 | First6ofFN + First6ofLN + email |
| Deterministic 3 | First6ofFN + First6ofLN + street_address |
| Probabilistic | FN, LN, Street address, Phone<br>• Blocking scheme 1: FN_Soundex + LN_Soundex<br>• Blocking scheme 2: First3ofFN + First3ofLN + state |

Abbreviations: FN, first name; LN, last name

The linkage methods were adopted from the existing method currently used by HDC and applied in a stepwise approach.[13] Linkages identified using deterministic method 1 were removed before deterministic method 2 was executed, and linkages identified using deterministic method 2 were removed before deterministic method 3 was executed (Table 1). Probabilistic blocking schemes 1 and 2 were sequentially executed after deterministic method 3. The threshold to declare a link was set to 90; therefore, linked pairs with a match score lower than 90 were not considered matches and labeled as “non-matches.”[13] A weight redistribution method was applied to the probabilistic linkage method in the presence of missing linkage data.[14]

PII did not leave the host servers and was deleted after the matching process was complete. Shared data were destroyed after the de-identified aggregate (i.e., total) linked-pair count was obtained. The linked-pair count was used to determine the go/no-go status of the larger CCR project.

## RESULTS

Aggregate data from the CSU-VTH and UCHealth are shown in **Table 2**. The most common species that visited the CSU-VTH were dogs (48,544), followed by horses (11,781) and cats (9,867). Females accounted for 47.9% of the animal patients and 53.4% of the human patients. There were more inpatient visits (16.5% versus 1.0%) and emergency visits (20.7% versus 7.8%) for animals than humans. There were 34,449 (out of 70,192 animals; 49%) animal patients and 2,425,388 (out of 3,234,314 people; 75%) human patients who had >1 healthcare encounter. The median time interval between first and last visits was shorter in animals (149 days) compared to humans (810 days). Venous blood draws were among the procedures common to both animals and humans, suggesting both datasets have robust laboratory measurements.

**Table 2:**
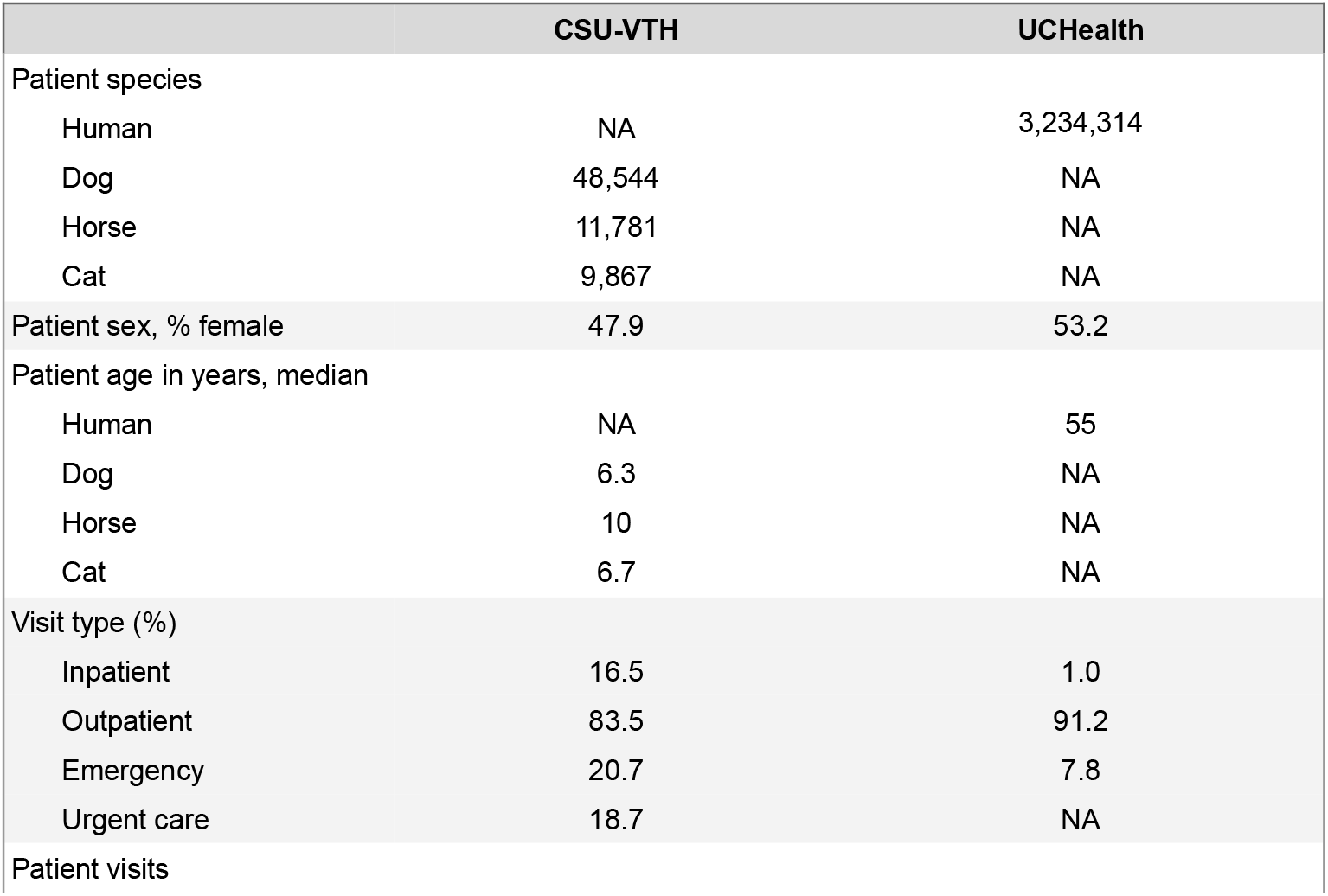
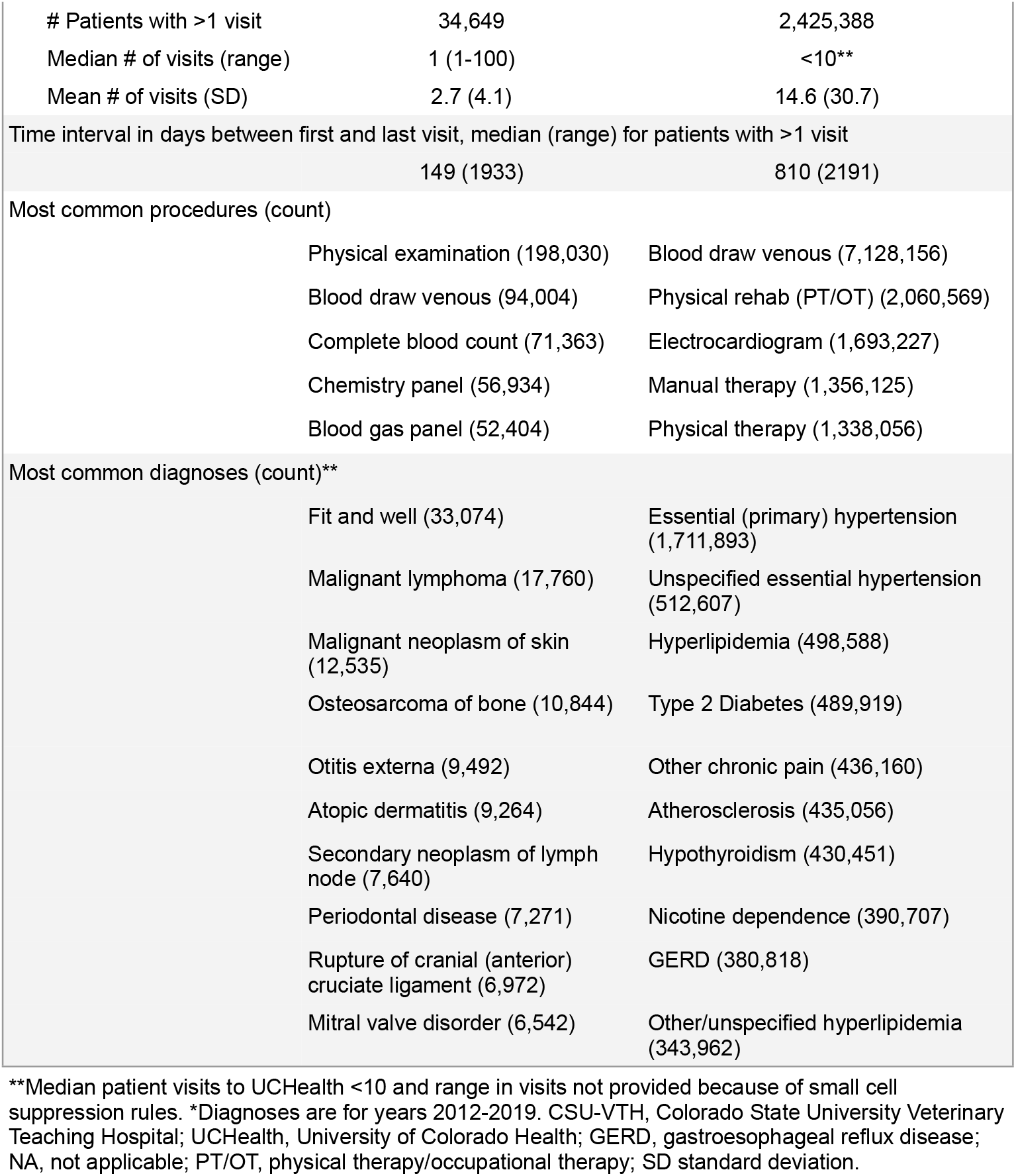
Characteristics of the CSU-VTH veterinary and UCHealth human patient populations respectively, 2019-2024.*

The most common diagnoses at the CSU-VTH were split between those typically seen in the primary care setting (i.e., fit and well (healthy), periodontal disease, otitis externa, and atopic dermatitis) and those seen in a veterinary referral hospital (i.e., malignant neoplasms, rupture of cranial (anterior) cruciate ligament, and mitral valve disorder).

The most common diagnoses from the UCHealth dataset generally represent chronic diseases (i.e., hypertension, hyperlipidemia, T2D, atherosclerosis, hypothyroidism, chronic pain, and gastroesophageal reflux disorder).

Attempts to evaluate comparative drug exposures were limited by non-standard categorization methods.

Securely linking non-medical PII from 41,081 CSU-VTH clients from August 2019-January 2024 with 3,282,860 UCHealth patients from 2015-2024 identified 12,115 matches, indicating that 29% of CSU-VTH clients were UCHealth patients. The CSU-VTH client population owned 76,282 animal patients: 55.5% were dogs, 15.3% were horses, 13.0% were cats, and 16.2% were other species (**Figure 1**).

**Figure 1:**
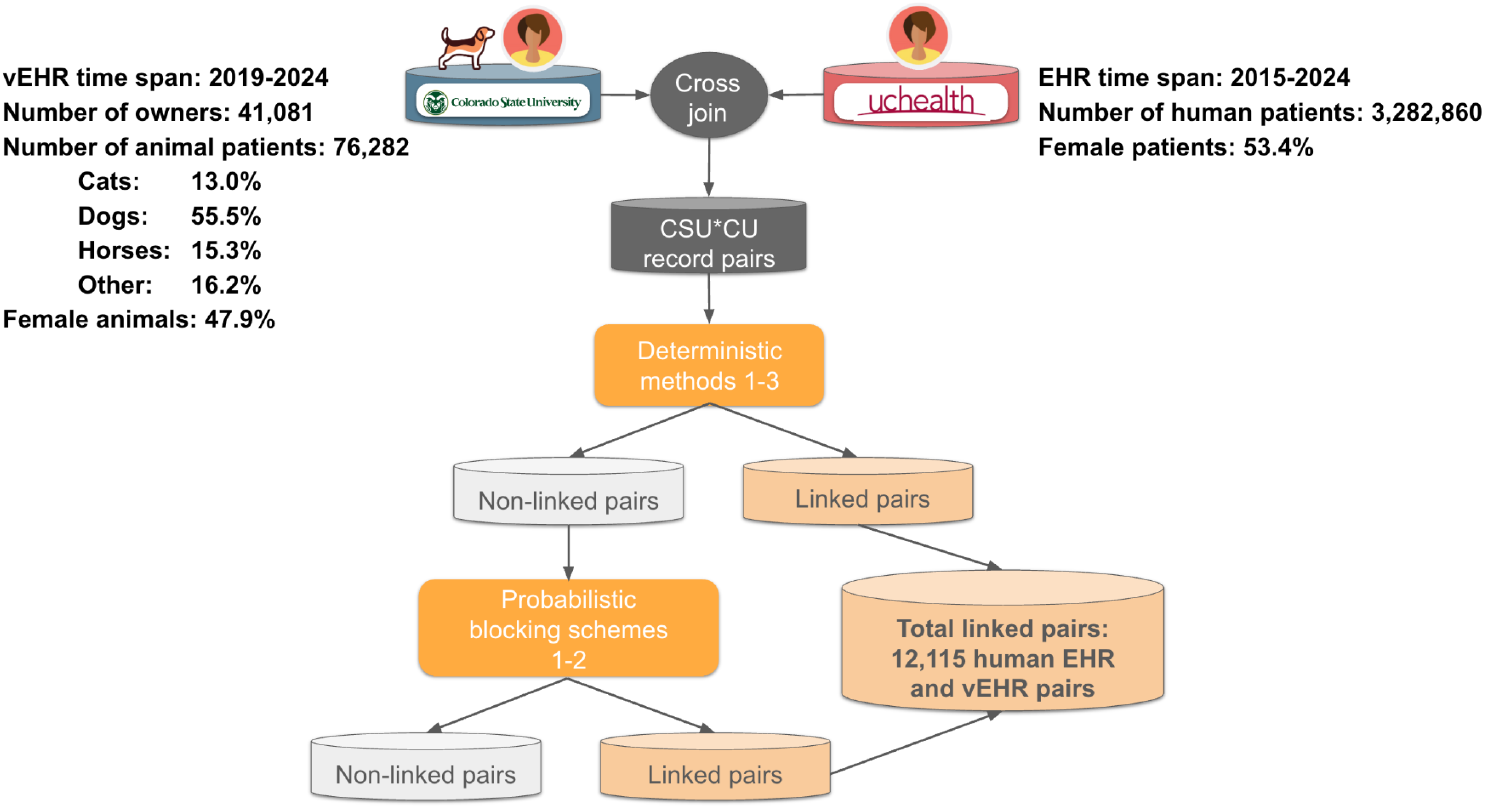
Non-medical Personally Identifying Information was linked from geographically adjacent human and veterinary medical academic institutions. The total number of matches (linked pairs of UCHealth patients who took their animal to the CSU-VTH) was 12,115, indicating 29% of companion animal owners were UCHealth patients. CSU-VTH, Colorado State University - Veterinary Teaching Hospital; EHR, electronic health record; UCHealth, University of Colorado Health; vEHR, veterinary electronic health record.

## DISCUSSION

This pilot study demonstrated the feasibility of developing a CCR utilizing EHR data for geographically-overlapping human and veterinary medical institutions. Comparison of the aggregate data across the institutions demonstrated similarities and differences between human and animal health and highlighted areas that need further standardization (e.g., drug categorization). Forty-nine percent of veterinary patients had greater than one healthcare encounter compared to 75% of human patients. This difference may be reflective of the CSU-VTH being a referral hospital, and future efforts to connect additional veterinary primary care clinics may be informative for the purpose of creating a longitudinal dataset of linked human-companion animal data.

Our successful bi-institutional data-sharing and patient matching identified 12,115 client-patient matches between the geographically adjacent hospitals. Considering the favorable results of this study, development of an OMOP[15–17] CCR for geographically overlapping human and veterinary medical institutions would enable investigation of the co-occurrence of conditions and environmental influences in coincident households. For example, several of the common diagnoses identified in the aggregate data from UCHealth, also occurred frequently in companion animals (e.g., diabetes, hypothyroidism, hypertension, and hyperlipidemia).[18–22] A CCR would facilitate evaluation of shared risk factors, etiologies, disease course, and treatment needs, options, and responses for these and other conditions.

Efforts to standardize data in vEHR systems using existing terminologies will improve the ability to use veterinary health data in comparative One Health efforts. Integrating open, community-driven, cross-species-enabled ontologies (e.g., from the Monarch Initiative[23,24]) into the CCR would allow more accurate capture of the rich phenotypic features of conditions and facilitate generalizability and interoperability between humans and veterinary species. Applying open, community-driven ontologies within the CCR will allow for integration of species-specific terms with precise mappings between terms allowing for cross-species comparisons.[25]

Creating the CCR requires strong collaborative efforts between institutions including attention to data governance and data management. Even with strong relationships, the current interinstitutional and interprofessional data sharing and feasibility study took significant time and effort to complete. Shared experiences and established collaborations, such as those created through the National Institutes of Health National Center for Advancing Translational Sciences Clinical and Translational Science Awards (CTSA), and commitment to One Health and interprofessional education efforts enable novel and integrated ways of finding solutions for current multi-species health concerns. For example, in a collaboration between Kansas State University and the University of Missouri, the 1DATA Project led to the development of a Master Sharing Agreement that serves as a template for expediting future One Health research.[26] In addition, the National COVID Cohort Collaborative (N3C) provides mechanisms and insights into creating a successful governance structure.[27] Key features that will be replicated in the CSU-CU CCR and across additional pairs of human and veterinary medical institutions within the CTSA One Health Alliance[28] include 1) obtaining institutional review board approval from each institution; 2) securing inter-institutional data sharing and use agreements; 3) engaging with veterinary stakeholders (pet owners, veterinarians, and veterinary administrators) to ensure transparency and that informed consent collected at admission reserves the right to use data in scientific studies 4) ensuring safe data transfer and storage from the veterinary hospital to the secure, HIPAA-compliant HDC RDW; 5) establishing a registry review board with members from the medical and veterinary institutions to review registry protocols, data access/use, training requirements, and analysis approvals within a tiered system (aggregate, de-identified, limited datasets); 6) ensuring the appropriateness of results output for dissemination; and 7) ensuring fair authorship and attribution.

Studies using data in the CCR will potentially lead to more involved data mining of vEHR data, sometimes in comparison to human records at the population level (e.g., comparing aspects of canine diabetes management with human diabetes management, irrespective of the household overlap), and sometimes in client-patient direct comparisons (e.g., exploring environmental factors affecting human and animal household health, such as household antimicrobial resistance patterns and the effects of wildfire smoke on incidence of household asthma).

The experiences and methods for designing the CCR are generalizable and will help overcome translational science barriers by leveraging cross-disciplinary research teams to advance whole-household health. By implementing the OMOP CDM, ensuring robust data privacy protocols, and establishing a comprehensive data governance framework, the registry will enable advanced cross-species analytical capabilities that when scaled up can support large-scale, cross-species analytics. We envision global network studies in One Health akin to what the OHDSI community is able to achieve.

## CONCLUSION

Data linkage across human and veterinary EHR is a feasible method to explore the dynamics of exposures and diseases shared by people and companion animals. The CCR will provide a blueprint for utilizing the OMOP CDM to facilitate future multi-species OHDSI network research studies, enabling deeper insights into human-animal-environment health interactions on a global scale.

## Data Availability

All data produced in the present work are contained in the manuscript

## Acknowledgments

The authors acknowledge the support of the Health Data Compass Data Warehouse project (healthdatacompass.org).

## Ethics Statement

Exemption status was obtained from the Colorado Multiple Institutional Review Board for the study (“Pilot Project to Assess Overlap between People Receiving Healthcare Locally (UCHealth) and Companion Animal Care at the CSU VTH” (COMIRB #23-0940)) and deemed not human subjects research by the Colorado State University Institutional Review Board (CSU IRB #1920).

## References

1. U.S. pet ownership statistics. American Veterinary Medical Association. https://www.avma.org/resources-tools/reports-statistics/us-pet-ownership-statistics (accessed 9 February 2025)

2. Facts + Statistics: Pet Ownership and Insurance. https://www.iii.org/fact-statistic/facts-statistics-pet-ownership-and-insurance (accessed 9 February 2025)

3. Mackenzie JS, Jeggo M. The One Health Approach-Why Is It So Important? Trop Med Infect Dis. 2019;4. doi: 10.3390/tropicalmed4020088

4. Linder DE, Santiago S, Halbreich ED. Is There a Correlation Between Dog Obesity and Human Obesity? Preliminary Findings of Overweight Status Among Dog Owners and Their Dogs. Front Vet Sci. 2021;8:654617.

5. Thielen SC, Reusch JEB, Regensteiner JG. A narrative review of exercise participation among adults with prediabetes or type 2 diabetes: barriers and solutions. Front Clin Diabetes Healthc. 2023;4:1218692.

6. Jagai JS, Krajewski AK, Shaikh S, et al. Association between environmental quality and diabetes in the USA. J Diabetes Investig. 2020;11:315–24.

7. Ruiz D, Becerra M, Jagai JS, et al. Disparities in Environmental Exposures to Endocrine-Disrupting Chemicals and Diabetes Risk in Vulnerable Populations. Diabetes Care. 2018;41:193–205.

8. Durward-Akhurst SA, Schultz NE, Norton EM, et al. Associations between endocrine disrupting chemicals and equine metabolic syndrome phenotypes. Chemosphere. 2019;218:652–61.

9. Iguacel I, Börnhorst C, Michels N, et al. Socioeconomically Disadvantaged Groups and Metabolic Syndrome in European Adolescents: The HELENA Study. J Adolesc Health. 2021;68:146–54.

10. Collins D, Lee H, Dunbar MD, et al. Associations between Neighborhood Disadvantage and Dog Walking among Participants in the Dog Aging Project. Int J Environ Res Public Health. 2022;19. doi: 10.3390/ijerph191811179

11. OHDSI – Observational Health Data Sciences and Informatics. https://ohdsi.org/ (accessed 12 February 2025)

12. Kahn MG, Mui JY, Ames MJ, et al. Migrating a research data warehouse to a public cloud: challenges and opportunities. J Am Med Inform Assoc. 2022;29:592–600.

13. Ong TC, Duca LM, Kahn MG, et al. A hybrid approach to record linkage using a combination of deterministic and probabilistic methodology. J Am Med Inform Assoc. 2020;27:505–13.

14. Ong TC, Mannino MV, Schilling LM, et al. Improving record linkage performance in the presence of missing linkage data. J Biomed Inform. 2014;52:43–54.

15. Liyanage H, Liaw S-T, Jonnagaddala J, et al. Common Data Models (CDMs) to Enhance International Big Data Analytics: A Diabetes Use Case to Compare Three CDMs. Stud Health Technol Inform. 2018;255:60–4.

16. Kwong M, Gardner HL, Dieterle N, et al. TRANSLATOR Database-A Vision for a Multi-Institutional Research Network. Top Companion Anim Med. 2019;37:100363.

17. Kwong M, Gardner HL, Dieterle N, et al. Optimization of Electronic Medical Records for Data Mining Using a Common Data Model. Top Companion Anim Med. 2019;37:100364.

18. Hoenig M. Comparative Aspects of Human, Canine, and Feline Obesity and Factors Predicting Progression to Diabetes. Veterinary Sciences. 2014;1:121–35.

19. Delicano RA, Hammar U, Egenvall A, et al. The shared risk of diabetes between dog and cat owners and their pets: register based cohort study. BMJ. 2020;371:m4337.

20. Tropf M, Nelson OL, Lee PM, et al. Cardiac and Metabolic Variables in Obese Dogs. J Vet Intern Med. 2017;31:1000–7.

21. McKenzie HC 3rd. Equine hyperlipidemias. Vet Clin North Am Equine Pract. 2011;27:59–72.

22. Lund EM, Jane Armstrong P, Kirk CA, et al. Prevalence and risk factors for obesity in adult dogs from private US veterinary practices. http://jarvm.com/articles/Vol4Iss2/Lund.pdf (accessed 13 November 2023)

23. Putman TE, Schaper K, Matentzoglu N, et al. The Monarch Initiative in 2024: an analytic platform integrating phenotypes, genes and diseases across species. Nucleic Acids Res. 2024;52:D938–49.

24. Callahan TJ, Stefanski AL, Wyrwa JM, et al. Ontologizing health systems data at scale: making translational discovery a reality. NPJ Digit Med. 2023;6:89.

25. Matentzoglu N, Bello SM, Stefancsik R, et al. The Unified Phenotype Ontology (uPheno): A framework for cross-species integrative phenomics. bioRxiv. 2024;2024.09.18.613276.

26. Staley J, Mazloom R, Lowe P, et al. Novel Data Sharing Agreement to Accelerate Big Data Translational Research Projects in the One Health Sphere. Top Companion Anim Med. 2019;37:100367.

27. Suver C, Harper J, Loomba J, et al. The N3C governance ecosystem: A model socio-technical partnership for the future of collaborative analytics at scale. J Clin Transl Sci. 2023;7:e252.

28. Lustgarten JL, Zehnder A, Shipman W, et al. Veterinary informatics: forging the future between veterinary medicine, human medicine, and One Health initiatives-a joint paper by the Association for Veterinary Informatics (AVI) and the CTSA One Health Alliance (COHA). JAMIA Open. 2020;3:306–17.

